# Aging-Related lncRNA Expression in Bipolar Disorder: Effects of Familial Liability and Childhood Trauma

**DOI:** 10.64898/2026.06.15.26355706

**Authors:** Sultan Ekinci, Buket Yeşiloğlu, İbrahim Fettahoğlu, Canay Pamukçu Ünlü, Hidayet Ece Arat Çelik, Şevin Hun Şenol, Sinem Balaç, Esma Çörekli Kaymakçı, Burcu Kök Kendirlioğlu, Mark A. Frye, Ayşegül Özerdem, Merih Altıntaş, Deniz Ceylan

## Abstract

**Introduction:** Bipolar disorder (BD) has been associated with increased medical burden and accelerated biological aging. Long non-coding RNAs (lncRNAs) regulate molecular pathways related to cellular senescence, inflammation, and telomere maintenance, which are implicated in both BD and aging. This study examined whether aging-related lncRNA expression reflects familial vulnerability or illness-specific effects, and whether childhood trauma and lifestyle factors modulate these signatures within a gene–environment framework.

**Methods:** In this cross-sectional study, expression levels of aging-related lncRNAs, including Antisense Non-coding RNA in the INK4 Locus (ANRIL), HOX Transcript Antisense Intergenic RNA (HOTAIR), Nuclear Enriched Abundant Transcript 1 (NEAT1), Taurine Upregulated Gene 1 (TUG1), Metastasis-Associated Lung Adenocarcinoma Transcript 1 (MALAT1), Growth Arrest-Specific 5 (GAS5), and Telomerase RNA Component (TERC), were measured in peripheral blood mononuclear cells (PBMCs) from individuals with bipolar disorder (BD) (n=68), siblings without BD diagnosis (SIB) (n=54), and healthy controls (HC) (n=70) using quantitative reverse transcription polymerase chain reaction (RT-qPCR). Childhood trauma and lifestyle were assessed using the Childhood Trauma Questionnaire (CTQ) and the Healthy Lifestyle Profile II (HPLP-II). Principal component analysis generated a composite aging-related lncRNA factor.

**Results:** At the individual transcript level, NEAT1 and TERC were elevated, whereas GAS5 was reduced, in both BD and SIB relative to HC. The aging-related lncRNA composite score was higher in BD and SIB than in HC (F = 7.315, p = 0.001). Familial liability to BD (presence vs. absence of familial liability) showed a significant main effect on the composite score (F(1,182)=8.18, p=0.005) and interacted with childhood trauma (F(1,182)=10.14, p=0.002). In multivariable models, total CTQ and physical neglect were independently associated with lower composite scores, while familial liability remained a positive predictor (all p<0.001).

**Conclusions:** Aging-related lncRNA alterations mark familial vulnerability to BD and are shaped by childhood trauma within a gene–environment interaction framework.

## Introduction

Bipolar disorder (BD) is a chronic, relapsing mood disorder characterized by manic/hypomanic and depressive episodes and is associated with substantial functional impairment, high cardiometabolic comorbidity, premature mortality, and accelerated biological aging [1, 2]. Despite its complex and still poorly understood underlying biology, several mechanisms have been consistently implicated in BD, including mitochondrial dysfunction, oxidative stress, and chronic low-grade inflammation, all of which overlap with central biological mechanisms of aging [3, 4]. Recent evidence suggests that these shared processes are at least partly mediated by epigenetic and post-translational regulatory pathways, including non-coding RNA-mediated regulation, whose dysregulation may provide a mechanistic link between aging biology and the pathophysiology of BD [5].

Long non-coding RNAs (lncRNAs) are regulatory RNA transcripts, exceeding 200 nucleotides in length, do not encode proteins but serve crucial regulatory functions in gene expression and cellular aging mechanisms [6]. In BD, altered expressions of lncRNAs involved in epigenetic regulation, mitochondrial function, and inflammatory pathways have been reported across multiple independent studies, supporting a role for lncRNA dysregulation in the molecular pathology of the disorder [7, 8, 9]. In aging-related diseases, or conditions in which advanced age constitutes an independent risk factor such as Alzheimer’s disease, cardiovascular disease, cancer, and neurodegenerative disorders, alterations in lncRNA expression have been reported to accompany age-dependent changes in epigenetic regulation, cellular senescence, mitochondrial function, and inflammatory signaling [10, 11]. Given their tissue specificity, stress responsiveness, and integrative role across genetic, epigenetic, and metabolic pathways, age-related lncRNAs represent compelling targets for investigation in BD.

Accumulating evidence implicates a subset of lncRNAs in cellular senescence, core aging-related mechanisms, and aging-associated neuropsychiatric disorders; antisense non-coding RNA in the INK4 locus (ANRIL), HOX transcript antisense intergenic RNA (HOTAIR), nuclear enriched abundant transcript 1 (NEAT1), taurine upregulated gene 1 (TUG1), metastasis-associated lung adenocarcinoma transcript 1 (MALAT1/NEAT2), and growth arrest-specific 5 (GAS5) have been implicated in both aging-related contexts and BD [5, 12–17]. However, their relevance to BD requires confirmation in independent, well-characterized cohorts, while other overlapping lncRNAs remain candidates for future investigation rather than established contributors to BD pathophysiology [9, 18]. Functionally, these lncRNAs regulate key aging-related pathways, including epigenetic repression via Polycomb repressive complexes (ANRIL, HOTAIR) [19–21], stress-responsive gene regulation and mitochondrial dysfunction (NEAT1), neurodegeneration [22, 23], mitochondrial bioenergetics and transcriptional homeostasis (TUG1, MALAT1), growth arrest and mTOR-dependent metabolic control (GAS5) [14, 15, 24]. In addition, although direct evidence demonstrating altered telomere-associated lncRNA TERC (telomerase RNA component) expression or regulation in BD is currently lacking, the extensive literature on telomere shortening in BD, together with the established roles of the telomerase RNA component TERC in telomere maintenance and genomic stability, supports the plausibility of telomerase-related dysregulation contributing to replicative senescence and premature cellular aging in BD [25–27]. Collectively, these aging-related lncRNAs may represent a molecular interface linking BD pathophysiology with biological pathways commonly involved in aging.

Given BD’s strong genetic basis, siblings without BD diagnosis (SIB) of individuals with BD often show overlapping biological alterations, suggesting trait-like markers of genetic liability [28, 29]. Genetic studies further demonstrate shared genetic loci between BD and cardiometabolic traits, linking genetic vulnerability to aging-related biological processes [30]. Importantly, these somatic conditions are not only more prevalent in individuals with BD but also significantly increased among their first-degree relatives [31, 32]. However, aging-related alterations in BD may also reflect environmental and illness-related exposures. Childhood trauma, a well-established risk factor for BD, and lifestyle-related factors have each been independently associated with accelerated biological aging and may confound observed aging signatures [33]. Therefore, examining SIB alongside patients and healthy controls (HC), while systematically assessing environmental and lifestyle exposures across all groups, is essential to disentangle genetic liability from environmental influences on aging-related vulnerability.

In this study, we assessed the expression profiles of seven lncRNAs identified in the literature as relevant to biological aging in BD across euthymic individuals with BD, SIB, and HC, and examined their associations with childhood trauma exposure and lifestyle-related factors. We hypothesized that aging-related lncRNA dysregulation would be present in both BD and SIB, consistent with familial vulnerability, and that environmental exposures would modulate these molecular patterns.

## Methods

### Study Design and Participants

This cross-sectional case-control study included 68 euthymic individuals with BD, 54 SIB, and 70 HC, all aged 18–50 years. Individuals with BD were receiving follow-up care at the Departments of Psychiatry of Maltepe University and Kartal Lütfi Kırdar City Hospital. Eligible SIB participants under active clinical follow-up were contacted by telephone and invited to participate, while HC participants were recruited from the surrounding community through informational brochures distributed in the hospital vicinity. All participants provided written informed consent prior to study procedures. The study protocol was approved by the Clinical Research Ethics Committee of Koç University (2024.182.IRB2.081; 2025.297.IRB2.138).

Psychiatric diagnoses, comorbid psychiatric conditions, and HC status were confirmed using the Structured Clinical Interview for DSM-5 (SCID-5). Exclusion criteria for all groups included decompensated medical conditions, morbid obesity, diabetes mellitus, rheumatologic diseases, active infections, neurological disorders, cognitive impairment, abnormal laboratory findings, risky alcohol use, pregnancy, menopause, breastfeeding, antioxidant supplementation, or substance use disorders (except tobacco). In the SIB group, individuals with BD, schizophrenia, or other psychotic disorders were excluded, whereas those with non-psychotic psychiatric comorbidities (e.g., depression, anxiety, ADHD) were retained due to their relevance to genetic liability for BD. In the HC group, any lifetime psychiatric diagnosis was an exclusion criterion. All participants met criteria for euthymia, defined as scores below 7 on both the 17-item Hamilton Depression Rating Scale (HDRS-17) [34, 35] and the Young Mania Rating Scale (YMRS) [36, 37]. Clinical assessments further included the self-report Health-Promoting Lifestyle Profile II (HPLP-II) [38, 39] and the Childhood Trauma Questionnaire (CTQ) [40, 41].

### RNA Extraction and Quantification of LncRNAs

Venous blood samples (10 mL) were collected between 09:00 and 10:00 AM after a 10-hour fast. Peripheral blood mononuclear cells (PBMCs) were then isolated using the density-gradient centrifugation method, and total RNA was extracted from PBMCs using the Qiagen RNeasy Mini Kit (Cat no./ID: 74104, Qiagen, Hilden, Germany), following the manufacturer’s instructions. The isolated RNA samples were stored at −80°C until further use. RNA concentration and purity were measured with a Thermo Scientific NanoDrop™ 2000 spectrophotometer using 260/280 and 260/230 ratios. RNA samples with low quality and/or insufficient concentration were excluded from further analysis. Complementary DNA (cDNA) synthesis was performed using the Bio-Rad iScript™ cDNA Synthesis Kit (Cat. No./ID: 1708890, Bio-Rad Laboratories, USA) in accordance with the manufacturer’s protocols.

Target lncRNAs were selected based on converging evidence from the literature, restricted to those implicated in both aging-related pathways and BD [5]. Primers for ANRIL, HOTAIR, NEAT1, TUG1, MALAT1, GAS5, TERC, and GAPDH were purchased from Oligomer Biotechnology (Istanbul, Türkiye). Primer sequences were obtained from previously published studies or, when necessary, redesigned using Primer3 software and are provided in Supplementary Table 1 [42]. Quantitative reverse transcription PCR (RT-qPCR) analyses were conducted in duplicate on a QuantStudio™ 7 Flex Real-Time PCR system at Koç University KUTTAM Laboratory, following MIQE guidelines [43]. Normalization of each sample set was performed using GAPDH as the reference gene in line with prior lncRNA expression studies using similar biological material and analytical approaches [44]. GAPDH was used as the reference gene; gene expression levels were normalized using the ΔCt method, and relative expression values were calculated using the 2^-ΔΔCt^ formula [45, 46].

### Statistical Analysis

Statistical analyses were performed using IBM SPSS Statistics version 29.0 (IBM Corp., Armonk, NY, USA, 2023) and RStudio version 2025.09.2+418 (Posit Software, Boston, MA, USA, 2025). Figures were generated using GraphPad Prism version 10.6.1 (GraphPad Software, San Diego, CA, USA, 2024) and RStudio. Categorical variables were compared using Chi-square tests. The Gaussian distribution of continuous data was assessed using the Kolmogorov-Smirnov test, Q-Q plots, and histograms, with data presented as means±standard deviations or medians with ranges, as appropriate. Demographic and clinical variables across study groups were compared using one-way analysis of variance (ANOVA) or Kruskal–Wallis tests for continuous variables, as appropriate, and Chi-square tests for categorical variables. For initial group comparisons of aging-related lncRNAs, Univariate Analyses of Covariance (ANCOVA) models were used when normality assumptions were met, adjusting for age and body mass index (BMI) as continuous covariates and sex and smoking status as categorical covariates. In the presence of non-Gaussian distributions, Quade’s Nonparametric Univariate Analyses of Covariance (Quade-ANCOVA) was used. A principal component analysis (PCA) was conducted to summarize shared variance across 7 aging-related lncRNAs. Sampling adequacy and factorability were confirmed using the Kaiser-Meyer-Olkin (KMO) measure and Bartlett’s test of sphericity. Accordingly, the seven lncRNAs were reduced to a single aging-related lncRNA composite score that captured their shared expression variance, as supported by the scree plot (Supplementary Figure S1). Oblimin rotation was used to allow for potential correlations between components. This aging-related lncRNA composite score was used in subsequent group comparisons and correlation analyses. The PCA approach also reduced dimensionality and helped control type I error due to multiple testing [47]. To meet normality assumptions, the composite score was log-transformed prior to analysis.

ANCOVA models were used to compare the lncRNA composite across groups, adjusting for age and BMI as continuous covariates, and for sex and smoking status as categorical covariates. Childhood trauma was modeled both as a continuous variable (total CTQ score) and as a binary variable (no exposure vs. exposure based on established cut-offs). To examine potential gene–environment interplay, interaction terms were incorporated into the models. Specifically, the interaction between binary familial liability to BD (presence vs. absence of familial liability) and binary childhood trauma exposure was tested by including a familial liability × CTQ (binary) term in the fully adjusted models. When statistically significant, simple effects were evaluated within strata defined by familial liability and trauma exposure. Correlation analyses were conducted in the total sample and separately within each diagnostic group using Spearman’s rank correlation coefficients for continuous variables. Point-biserial correlation analyses were used to evaluate associations between dependent variables and categorical clinical variables (e.g., sex, smoking status, medication use). Correlation strength was interpreted as moderate for |r|≥0.40. Multiple linear regression analyses were conducted to identify predictors of the aging-related lncRNA composite score.

Clinical variables that showed significant correlations with the composite score were included as predictors, within adjustments for sex, age, BMI, and smoking status. Separate regression models were also fitted for each group to examine group-specific associations. In stratified multiple linear regression analyses, childhood trauma subtypes were entered as binary variables (presence vs. absence) based on established CTQ cutoff scores, with the aging-related lncRNA composite as the dependent variable, adjusting for age, sex, smoking status, and body mass index. Statistical significance was set at p<0.05 for all analyses, p-values were adjusted for multiple comparisons using the Benjamini–Hochberg (BH) procedure with FDR=0.05 [48].

## Results

### Sociodemographic and clinical characteristics

The demographic and clinical characteristics of all groups are summarized in Table 1. There were no significant group differences in sex distribution, age, years of education, or smoking status. In contrast, significant differences were observed in partnership status (p=0.002), employment status (p<0.001), and BMI (p<0.001), with individuals with BD showing lower rates of partnership and employment and higher BMI compared to the other groups (Table 1).

**Table 1.** Sociodemographic and clinical characteristics of the study groups.

| Variable | BD (n=68)<br>Mean ± SD /<br>% | SIB (n=54)<br>Mean ± SD /<br>% | HC (n=70)<br>Mean ± SD /<br>% | F / $\chi^2$ | df | p |
| --- | --- | --- | --- | --- | --- | --- |
| Age (years) | 35.41 ± 7.24 | 36.48 ± 7.93 | 33.53 ± 7.03 | 2.59 | 2 | ns |
| Sex (female %) | 67.20% | 63.00% | 52.90% | 3.1 | 2 | ns |
| Partnership<br>status (with<br>partner %) | 29.20% | 55.60% | 57.10% | 17.29 | 2 | 0.002 |
| Working status<br>(employed %) | 53.80% | 74.10% | 97.10% | 34.41 | 2 | <0.001 |
| Years of<br>education | 13.89 ± 3.09 | 14.02 ± 3.90 | 14.76 ± 2.56 | 1.45 | 2 | ns |
| BMI (kg/m<sup>2</sup>) | 27.96 ± 5.68 | 24.55 ± 3.57 | 24.48 ± 3.32 | 13.47 | 2 | <0.001 |
| Medication use<br>(%) | 97.00% | 13.00% | 0% | 156.8 | 2 | <0.001 |
| Smoking status<br>(non-smoker %) | 43.90% | 55.60% | 47.10% | 1.68 | 2 | ns |
| Childhood<br>trauma (yes%) | 64.7% | 57.4% | 24.3% | 25.292 | 2 | <0.001 |
| HDRS-17 | 1.85 ± 1.57 | 1.02 ± 1.24 | 1.05 ± 1.52 | 6.41 | 2 | 0.002 |
| YMRS | 0.50 ± 0.96 | 0.08 ± 0.34 | 0.05 ± 0.22 | 10.22 | 2 | <0.001 |
| HPLP-II Total | 123.21 ±<br>19.88 | 131.84 ± 16.21 | 143.28 ± 21.83 | 17.9 | 2 | <0.001 |
| Health<br>responsibility | 21.26 ± 4.45 | 21.17 ± 4.07 | 23.54 ± 5.21 | 5.55 | 2 | 0.005 |
| Physical activity | 15.07 ± 5.08 | 17.12 ± 4.80 | 19.86 ± 5.82 | 14.11 | 2 | <0.001 |
| Nutrition | 19.07 ± 4.35 | 20.32 ± 3.36 | 21.35 ± 3.75 | 5.94 | 2 | 0.003 |
| Spiritual growth | 24.55 ± 4.53 | 26.72 ± 4.26 | 28.33 ± 3.56 | 14.46 | 2 | <0.001 |
| Interpersonal relations | 24.01 ± 4.22 | 26.07 ± 4.10 | 28.16 ± 3.45 | 19.18 | 2 | <0.001 |
| Stress management | 19.25 ± 3.74 | 20.44 ± 3.67 | 22.04 ± 4.48 | 8.35 | 2 | <0.001 |
| **CTQ-Total** | 47.53 ± 17.33 | 41.89 ± 12.95 | 35.32 ± 8.91 | 14.111 | 2 | <0.001 |
| CTQ – Emotional abuse | 9.89 ± 4.47 | 8.35 ± 3.53 | 6.45 ± 2.61 | 15.58 | 2 | <0.001 |
| CTQ – Physical abuse | 7.30 ± 3.59 | 6.33 ± 2.46 | 5.49 ± 1.85 | 7.53 | 2 | <0.001 |
| CTQ – Physical neglect | 10.32 ± 4.26 | 8.87 ± 3.99 | 9.08 ± 4.12 | 2.29 | 2 | ns |
| CTQ – Sexual abuse | 7.34 ± 4.88 | 6.03 ± 3.04 | 5.27 ± 0.84 | 6.63 | 2 | 0.002 |

### LncRNA expression levels

NEAT1 and TERC expression levels were significantly higher in BD and SIB compared with HC, whereas GAS5 levels were significantly lower in both BD and SIB than in HC. No significant differences were observed between BD and SIB for these transcripts. TUG1 and MALAT1 also showed significant group differences, both driven by higher expression in SIB. ANRIL and HOTAIR did not differ significantly across groups (Table 2, Figure 1).

**Figure 1.**
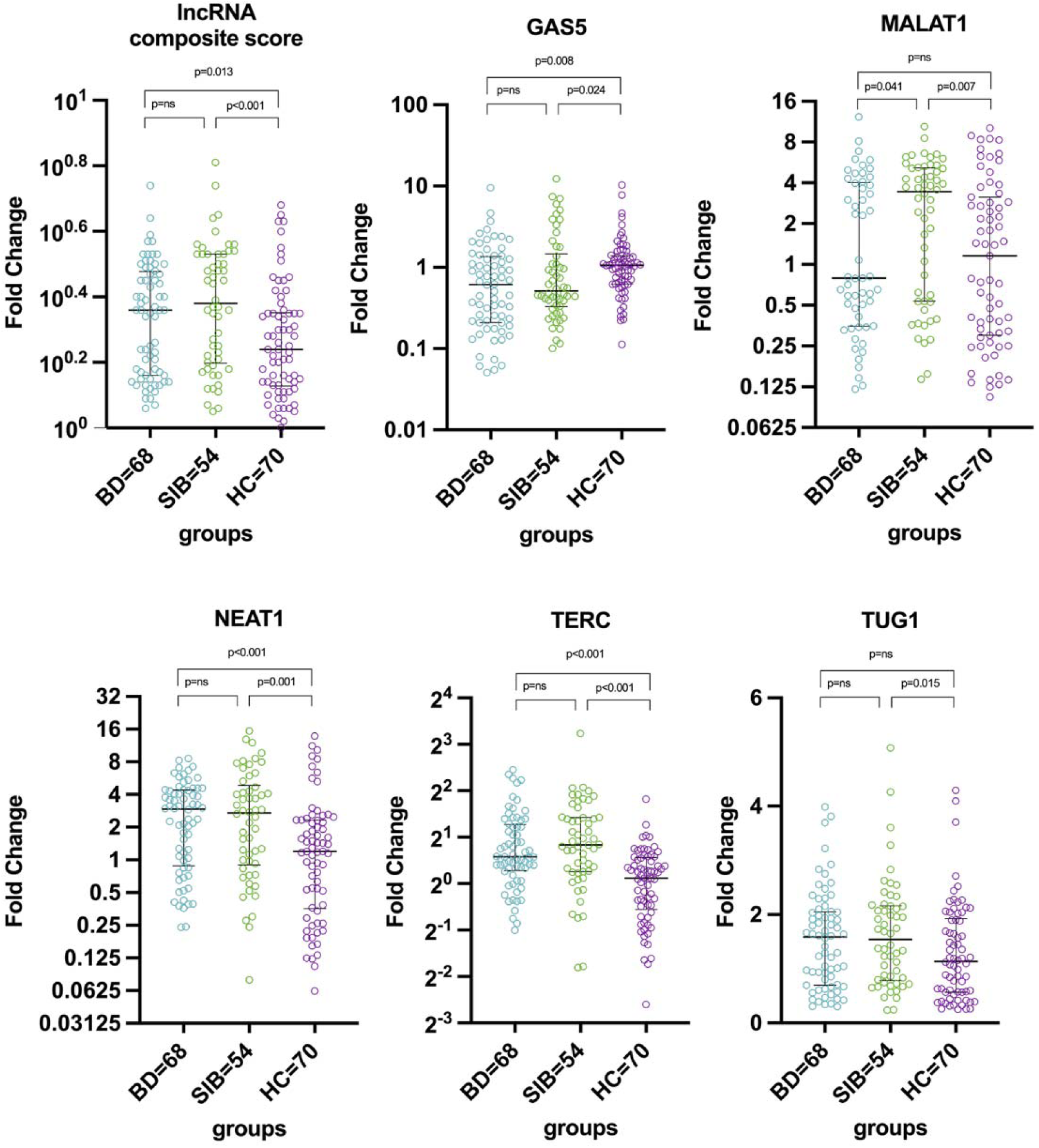
Expression levels of aging-related lncRNAs across study groups. Expression levels of ANRIL, HOTAIR, NEAT1, TUG1, MALAT1, GAS5, and TERC in individuals with BD, SIB, and HC. Individual data points are displayed together with group medians and interquartile ranges. Statistical comparisons were performed using ANCOVA models adjusted for age, sex, body mass index, and smoking status.

**Table 2.** Comparison of lncRNA expression levels among study groups.

| **lncRNA**<br>**(2<sup>-</sup>ΔΔCt)** | **BD**<br>**Median (min–**<br>**max)**<br>**Mean ± SD** | **SIB**<br>**Median (min–**<br>**max)**<br>**Mean ± SD** | **HC**<br>**Median (min–**<br>**max)**<br>**Mean ± SD** | **F**<br>**(DFH,**<br>**DFE)** | **p** | **Post-hoc**<br>**comparisons**<br>**(p < 0.05)** |
| --- | --- | --- | --- | --- | --- | --- |
| **NEAT1** | 2.90 (0.24–8.58)<br>2.87 ± 2.11 | 2.70 (0.08–<br>15.44)<br>3.59 ± 3.51 | 1.20 (0.06–<br>13.82)<br>2.10 ± 2.81 | 8.674<br>(2,185) | <0.001 | BD > HC<br>SIB > HC |
| **TERC** | 1.49 (0.50–5.46)<br>1.85 ± 1.07 | 1.79 (0.29–9.41)<br>2.15 ± 1.46 | 1.11 (0.16–5.83)<br>1.18 ± 0.80 | 17.298<br>(2,185) | <0.001 | BD > HC<br>SIB > HC |
| **TUG1** | 1.57 (0.30–3.99)<br>1.50 ± 0.87 | 1.54 (0.24–5.08)<br>1.64 ± 1.00 | 1.14 (0.25–4.29)<br>1.30 ± 0.91 | 3.466<br>(2,185) | 0.033 | SIB > HC |
| **MALAT1** | 0.79 (0.12–8.13)<br>2.07 ± 2.15 | 3.44 (0.14–<br>10.43)<br>3.20 ± 2.49 | 1.16 (0.10–<br>10.17)<br>2.25 ± 2.64 | 3.981<br>(2,168) | 0.020 | SIB > BD<br>SIB > HC |
| **GAS5** | 0.61 (0.05–9.50)<br>1.01 ± 1.40 | 0.51 (0.10–<br>12.22)<br>1.52 ± 2.31 | 1.06 (0.11–<br>10.26)<br>1.38 ± 1.57 | 4.334<br>(2,185) | 0.014 | BD < HC<br>SIB < HC |
| **ANRIL** | 0.87 (0.30–3.84)<br>1.10 ± 0.73 | 1.13 (0.32–<br>45.21)<br>2.11 ± 6.02 | 1.02 (0.20–8.07)<br>1.27 ± 1.10 | 1.375<br>(2,185) | ns | ns |
| **HOTAIR** | 1.09 (0.01–<br>167.69)<br>7.35 ± 24.17 | 1.02 (0.02–<br>128.35)<br>6.53 ± 19.36 | 1.55 (0.01–<br>29.89)<br>4.22 ± 7.40 | 0.013<br>(2,152) | ns | ns |
| **Aging-**<br>**related**<br>**lncRNA**<br>**composite** | 0.36 (0.06-0.64)<br>0.32±0.16 | 0.39 (0.05-0.81)<br>0.38± 0.19 | 0.24 (0.00-0.68)<br>0.26±0.17 | 7.315<br>(2,185) | 0.001 | BD > HC<br>SIB > HC |

### Aging-related lncRNA composite score

Sampling adequacy was assessed using the Kaiser–Meyer–Olkin (KMO) measure, which indicated acceptable suitability for factor analysis (KMO=0.602). Bartlett’s test of sphericity confirmed that the correlation matrix was appropriate for component extraction (χ²=493.905, df=21, p<0.001). Components with eigenvalues greater than 1 were retained. Eigenvalues > 1 and the inspection of the scree plot both supported a single-factor solution. PCA results indicated that the first component had an eigenvalue of 2.824, accounting for 40.3% of the total variance. The second and third components had eigenvalues of 1.418 and 1.117, explaining 20.3% and 16.0% of the variance, respectively, while subsequent components accounted for less than 11% each. Although three components exceeded the eigenvalue >1 criterion, inspection of the scree plot suggested a dominant primary component. The aging-related lncRNA composite score was significantly higher in BD (0.36 [0.06–0.64]) and SIB (0.39 [0.05–0.81]) than in HC (0.24 [0.00–0.68]) (F = 7.315; BD vs. HC: t = 2.496, p = 0.013; SIB vs. HC: t = 3.842, p < 0.001), with no difference between BD and SIB (Figure 1).

### Interaction between familial liability and childhood trauma

In the ANCOVA model predicting lncRNA composite, a significant interaction between childhood trauma and familial liability was observed (β=−0.18, p=0.002), after adjustment for age, sex, smoking status and BMI, with no other interactions (Figure 2). In contrast, the main effect of childhood trauma alone was not significant (F(1,182)=0.83, p=0.365). The three-way interaction between childhood trauma, familial liability, and sex did not reach statistical significance (F(4,182)=1.77, p=0.138). The overall model was significant (R²=0.168, adjusted R²=0.136, p<0.001).

**Figure 2.**
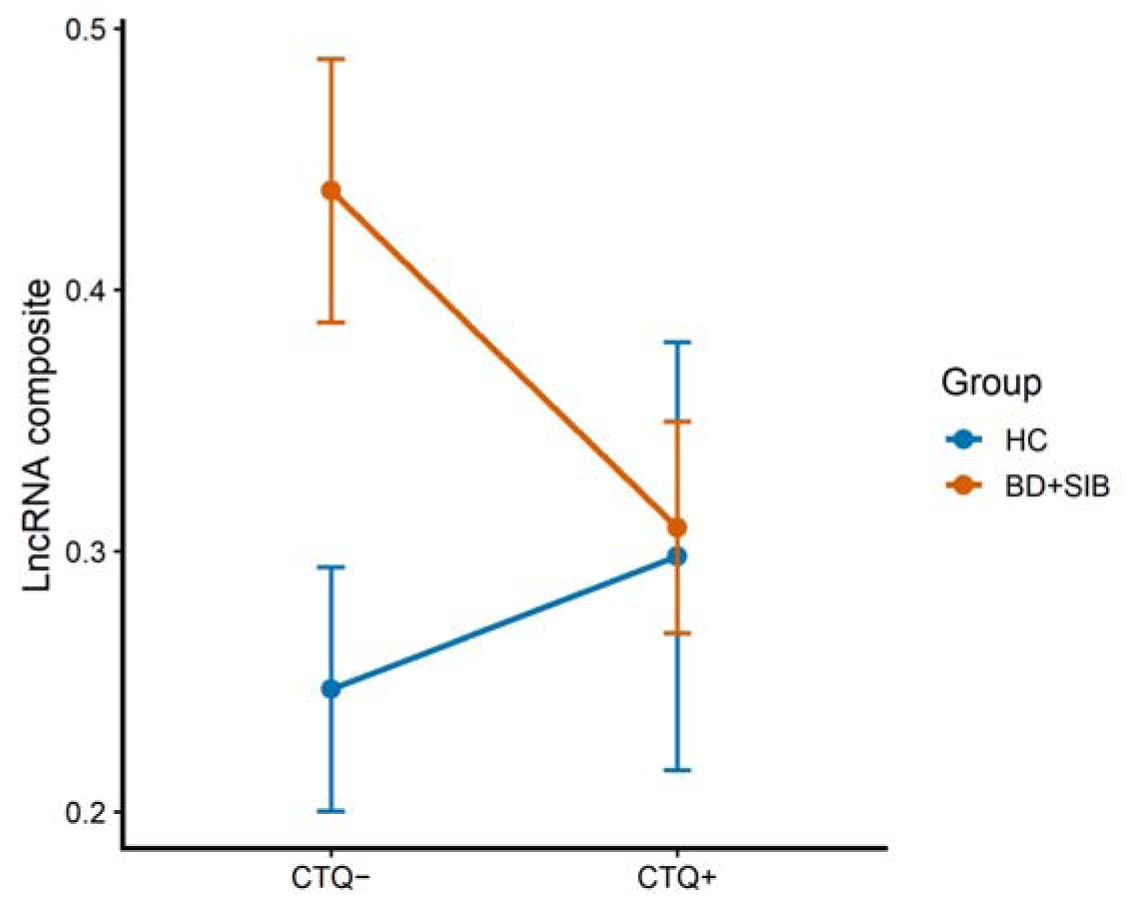
Interaction between childhood trauma and familial liability on the aging-related lncRNA composite score. Adjusted estimated marginal means of the lncRNA composite score according to childhood trauma exposure (CTQ− vs CTQ+) and familial liability status (liability− vs liability+), controlling for age, sex, smoking status, and body mass index. A significant interaction between familial liability and childhood trauma was observed.

### Aging-related lncRNA composite and clinical associations

In the total sample, no correlations exceeded the predefined threshold of |r| ≥ 0.40. However, modest but statistically significant negative correlations were observed between the aging-related lncRNA composite score and both depressive symptom severity (HDRS-17 scores) and the number of depressive episodes (r = −0.394, p < 0.001; r = −0.385, p < 0.001). In group-specific analyses, moderate to strong correlations were observed. In the BD group, the composite score was negatively associated with HDRS-17 scores (r = −0.513, p < 0.001) and childhood physical neglect (r = −0.697, p < 0.001), and positively associated with the number of depressive episodes (r = 0.473, p < 0.001). In the SIB group, the composite score was negatively associated with childhood physical neglect (r = −0.588, p < 0.001), physical abuse (r = −0.410, p = 0.002), and total CTQ score (r = −0.443, p < 0.001). In the HC group, the composite score was negatively associated with HDRS-17 scores (r = −0.444, p < 0.001). No correlations exceeding |r| ≥ 0.40 were observed for lifestyle factors or other demographic or clinical variables.

In multivariable linear regression analyses, in the total sample, adjusted for age, sex, smoking status, and body mass index, higher total CTQ scores were independently associated with lower aging-related lncRNA composite scores (B=−0.004, 95% CI −0.006 to −0.002, p<0.001). Familial liability status (BD or SIB vs. HC) was positively associated with the aging-related lncRNA composite (B=0.144, 95% CI 0.081 to 0.206, p<0.001), whereas clinical illness status (BD vs. SIB or HC) was not independently associated. In a separate model focusing on childhood physical neglect, physical neglect was independently associated with lower aging-related lncRNA composite levels (B=−0.020, β=−0.460, 95% CI −0.026 to −0.014, p<0.001). The overall model was significant (R²=0.212, F(5,184) =9.88, p<0.001), while other covariates were not significant.

In stratified analyses using nominal childhood trauma variables defined by established CTQ cutoff scores (presence vs. absence), separate multiple linear regression models were fitted for participants without familial liability (i.e., HC) and those with familial liability (i.e., BD and SIB). In the HC group, the overall model was significant (R²=0.207, F(9,58)=2.94, p=0.006). Childhood physical neglect was negatively associated with the aging-related lncRNA composite (B=−0.075, β=−0.221, 95% CI −0.149 to 0.000, p=0.049), while childhood sexual abuse was positively associated (B=0.231, β=0.441, 95% CI 0.104 to 0.358, p<0.001). In the familial liability group (i.e., BD and SIB), the regression model was also significant (R²=0.224, F(9,106)=4.70, p<0.001), with childhood physical neglect emerging as the only significant predictor of the aging-related lncRNA composite (B=−0.170, β=−0.472, 95% CI −0.240 to −0.099, p<0.001). No other childhood trauma subtype or covariate reached statistical significance.

## Discussion

In this cross-sectional study of euthymic individuals with BD, SIB, and HC, the principal finding was that an aging-related lncRNA composite score was higher in both BD and SIB than in HC, whereas BD and SIB did not differ from each other. In multivariable analyses, familial liability to BD, rather than diagnosis alone, was associated with the composite score. Additionally, childhood trauma exposure, particularly physical neglect, was associated with lower composite levels, and this association varied according to familial liability. Taken together, these findings suggest that aging-related lncRNA dysregulation in BD may be more closely linked to familial vulnerability and may be modified by early environmental adversity.

To contextualize these results, we examined aging-associated lncRNAs involved in cellular senescence, stress responsiveness, mitochondrial function, and genomic stability. In our cohort, NEAT1 expression was significantly higher in individuals with BD and in SIB compared with HC, whereas no difference was observed between the BD and SIB groups. TERC expression was also increased in both BD and SIB relative to HC, consistent with potential involvement of telomere-related regulatory mechanisms in aging processes associated with genetic vulnerability. GAS5 expression was reduced in both BD and SIB compared with HC. In contrast, no significant BD–HC differences were observed for TUG1, MALAT1, ANRIL, or HOTAIR, suggesting that alterations in these lncRNAs may be context-dependent or reflect factors beyond the core clinical phenotype of BD.

Previous studies have reported heterogeneous expression patterns of aging-related lncRNAs in BD, including largely null findings for NEAT1 [49], inconsistent results for TUG1 and MALAT1 [7, 50–52], relatively consistent downregulation of GAS5 [24, 53], and no expression-level differences for HOTAIR or ANRIL [19–21]. In this context, our observation of increased NEAT1 expression in BD contrasts with earlier null findings and extends the literature by demonstrating a parallel elevation in SIB, supporting a potential trait-like association with genetic vulnerability. The reduction of GAS5 in BD is consistent with prior reports, reinforcing its role as a stress– and aging-related vulnerability marker. Conversely, the absence of BD–HC differences for TUG1 and MALAT1 in our data mirrors the variability reported in the literature, supporting state– or context-dependent regulation rather than stable disease markers. Consistent with previous studies, we observed no significant expression differences for ANRIL or HOTAIR.

Critically, our findings are strengthened by replication in SIB, who share genetic vulnerability but lack illness-related confounds such as medication effects. NEAT1, TERC, and GAS5 showed consistent directional changes in both BD and SIB relative to HC, supporting the possibility that these lncRNAs reflect trait-like familial vulnerability rather than illness-state markers. This convergence across BD and SIB groups is consistent with a trait-related pattern suggesting that these molecular signatures may reflect inherited vulnerability to accelerated aging processes in BD, independent of clinical status. TUG1 showed sibling-specific elevation, potentially reflecting adaptive mitochondrial responses via AMPK–SIRT1 signaling and regulation of mitophagy pathways [54, 55]. Most notably, MALAT1 expression was elevated specifically in SIB but not in BD, suggesting a potential compensatory or resilience-related response in individuals with familial liability who remain clinically unaffected. Given MALAT1’s dynamic regulation by oxidative stress and metabolic state [7, 50, 56], this elevation may reflect adaptive or resilience-related mechanisms buffering cellular stress associated with inherited vulnerability.

Across analyses, correlations between the aging-related lncRNA composite and clinical or environmental variables were generally weak in the total sample, likely reflecting substantial biological heterogeneity. In HC, both aging-related lncRNA expression and CTQ scores were low and narrowly distributed, which may have limited statistical power to detect associations. In contrast, group-specific analyses revealed a clear divergence: individuals with familial liability to BD, including both affected individuals and their SIB, showed similar association patterns that differed from HC, suggesting that the aging-related lncRNA profile is more closely linked to genetic vulnerability than to diagnostic status alone.

Multivariable regression analyses, which were more informative than simple correlations, identified childhood trauma burden, particularly childhood physical neglect, as the strongest independent predictor of lower aging-related lncRNA composite levels after covariate adjustment. Familial liability status was independently associated with higher lncRNA levels, whereas clinical illness status was not, indicating opposing main effects of genetic vulnerability and early-life adversity. Importantly, ANCOVA analyses further demonstrated a significant interaction between familial liability and childhood trauma, indicating that the impact of childhood trauma on aging-related lncRNA expression differs according to familial liability status. This interaction suggests a gene–environment interplay rather than purely additive effects and highlights etiological heterogeneity in the biological embedding of early-life adversity.

Taken together, the differential expression patterns across these lncRNAs indicate that aging-related molecular alterations in BD are heterogeneous and context-dependent, comprising both relatively stable markers of inherited vulnerability (e.g., NEAT1, TERC, GAS5) and more dynamic, potentially adaptive regulators (e.g., TUG1, MALAT1). This gene-specific dissociation suggests that aging-related lncRNA dysregulation in BD should not be viewed as a uniform signature of accelerated biological aging, but rather as the convergence of partially overlapping molecular pathways shaped by genetic liability and environmental context. Consistent with emerging models of BD heterogeneity, these findings imply that genetically driven aging-related mechanisms may predominate in some individuals, whereas environmental stress–related pathways may be more influential in others. The absence of comparable trauma-related effects in HC further supports the notion that genetic vulnerability provides a permissive background upon which early-life adversity shapes aging-related molecular trajectories, underscoring the need to integrate genetic and environmental factors when interpreting molecular aging signatures in BD. Consistent with prior evidence of molecular alterations in SIB of individuals with BD, the inclusion of SIB in our study enabled us to distinguish illness-related lncRNA changes from genetically driven aging-associated molecular endophenotypes [57].

Notably, we observed a robust association between childhood trauma exposure, particularly physical neglect, and aging-related lncRNA expression within genetically vulnerable groups, with higher CTQ burden associated with lower lncRNA levels despite overall elevation in BD and SIB. One plausible explanation for this pattern is the marked genetic heterogeneity of BD, whereby a subset of individuals may carry a higher intrinsic genetic load directly linked to elevated lncRNA expression and aging-related pathways, while BD arising predominantly in the context of severe childhood trauma may reflect a partially distinct etiological trajectory with attenuated or differently patterned lncRNA alterations. This interpretation is supported by the inverse association between CTQ burden and lncRNA levels within the familial liability groups despite higher overall lncRNA expression and is consistent with prior BD literature showing that certain comorbidities, such as alcohol or cannabis use disorders, may index etiologically distinct subgroups rather than greater disease severity [58,59]. Collectively, these findings suggest that the aging-related lncRNA composite reflects a biologically meaningful dimension of BD liability shaped by the balance between genetic predisposition and early-life environmental stress, rather than diagnostic status alone.

On the other hand, no significant associations were observed between the aging-related lncRNA composite and lifestyle-related factors in either HC or BD, suggesting that these expression patterns may be less influenced by current behavioral exposures and instead reflect the enduring impact of earlier-life biological and environmental influences on gene regulation. This null finding should also be interpreted in the context of the study’s age range, which was restricted to adulthood and excluded individuals older than 50 years in line with the BD aging literature [60]. It is therefore possible that cumulative lifestyle effects on aging-related lncRNA expression become more apparent later in life and could not be captured within the present age window. Future studies that include older age groups and longer exposure histories will be needed to clarify the role of lifestyle-related influences as age advances.

One of the main strengths of this study is its comparative design, which allowed aging-associated lncRNA expression to be examined across three carefully characterized groups. Including SIB provided a valuable opportunity to distinguish biological patterns associated with genetic liability from those associated with the disorder’s clinical manifestation. Another important strength is the focus on a panel of lncRNAs previously implicated in aging-related processes, and their evaluation both individually and as a composite index, which offers a broader perspective on molecular mechanisms linked to biological aging. The study also benefited from the simultaneous consideration of key environmental and biological factors, including childhood trauma, familial liability status, and sex, within multivariable models. This approach made it possible to explore potential interactions between inherited vulnerability and early-life adversity. In addition, the inclusion of euthymic patients reduced the likelihood that acute mood symptoms influenced the observed molecular findings, thereby supporting the interpretation of more stable, trait-related biological characteristics.

Several limitations should be considered. First, the cross-sectional design precludes causal inference and limits conclusions about the temporal relationship between trauma exposure, genetic liability, and lncRNA alterations; longitudinal follow-up of patients and SIB is needed. Second, although major current mood episodes and psychotic disorders were excluded, some SIB had a history of other psychiatric conditions, which may have introduced heterogeneity. Third, lncRNA expression was measured in peripheral blood, which may not fully reflect brain-specific molecular processes; therefore, the findings should be interpreted cautiously. Fourth, the inclusion of only euthymic BD patients under age 50 limits generalizability to acute mood states and later-life aging. Finally, despite adjustment for major confounders, residual effects of illness duration, medication exposure, comorbidities, and lifestyle factors cannot be ruled out.

In conclusion, this study shows that aging-related lncRNA expression is altered in individuals with BD and their SIB, suggesting a stronger association with genetic vulnerability than with clinical illness status alone. Early-life adversity, particularly childhood trauma, was associated with reduced lncRNA expression within the familial liability group, suggesting that environmental exposures shape these molecular profiles. Overall, the findings indicate that aging-related lncRNAs may represent a biological interface between genetic predisposition and environmental stress, contributing to the heterogeneity and underlying vulnerability in BD.

## Funding

This research was funded by Koç University Faculty of Medicine.

## Conflict of Interest

The authors declare that they have no competing financial interests related to this work.

## Data Availability

Due to privacy and ethical restrictions involving human participant data, datasets are available from the corresponding author upon reasonable request.

## Acknowledgements

The authors thank all participants who took part in this study. The authors also acknowledge the technical support provided by the Koç University Center for Translational Medicine (KUTTAM) laboratory staff.

## Notes

### Competing Interest Statement

The authors have declared no competing interest.

### Author Declarations

The study protocol was approved by the Clinical Research Ethics Committee of Koc University (2024.182.IRB2.081; 2025.297.IRB2.138).

